# The Epidemiology Workbench: a Tool for Communities to Strategize in Response to COVID-19 and other Infectious Diseases

**DOI:** 10.1101/2020.07.22.20159798

**Authors:** Santiago Núñez-Corrales, Eric Jakobsson

**Affiliations:** Informatics, NCSA and MCB, UIUC

## Abstract

COVID-19 poses a dramatic challenge to health, community life, and the economy of communities across the world. While the properties of the virus are similar from place to place, the impact has been dramatically different from place to place, due to such factors as population density, mobility, age distribution, etc. Thus, optimum testing and social distancing strategies may also be different from place to place. The Epidemiology Workbench provides access to an agent-based model in which a community’s demographic, geographic, and public health information together with a social distancing and testing strategy may be input, and a range of possible outcomes computed, to inform local authorities on coping strategies. The model is adaptable to other infectious diseases, and to other strains of coronavirus. The tool is illustrated by scenarios for the cities of Urbana and Champaign, Illinois, the home of the University of Illinois at Urbana-Champaign. Our calculations suggest that massive testing is the most effective strategy to combat the likely increase in local cases due to mass ingress of a student population carrying a higher viral load than that currently present in the community.

## 1 Introduction

Mathematical models of infectious disease epidemiological dynamics can be provide valuable assistance to public health officials and health care providers in assessing the likely seriousness of an epidemic or its potential to grow into a pandemic, and later in allocating resources to counter the spread of the disease [105]. Simulations that trace either prior or projected time courses make use of various mathematical and computational techniques, including (non-exhaustively) differential equation models, statistical regression and curve fitting, network propagation dynamics, and direct representation of human actors and their actions by means of agent-based models. The SIR model in particular along with its various adaptations [100] has remained successful, at least in part, due to its universality as evidenced by its empirical adequacy across multiple epidemics, and by its formal robustness when connecting microscale host-pathogen related events and macroscale disease observables [28, 10].

Stochastic versions of the SIR model show that adding noise to the system changes the predicted onset of an epidemic [113], the stability of its endemic equilibrium [123], the value of its effective reproduction number [57] or its duration [72] when contrasted against the deterministic one. This is significant not only at the theoretical level when studying the stability and asymptotic representativeness of deterministic vs stochastic SIR models under various noise regimes, but at the policy making level where computational epidemiology may form the basis of informed decisions under policy, budgetary and other types of constraints [46].

Moreover, the SIR has been extended spatially to account for the diffusion-like properties associated with geographic patterns observed during epidemics. While the qualitative (and some quantitative) properties of the traditional and the spatial SIR models remain largely shared, spatial versions appear to be numerically susceptible to how these capture spatial interactions [101]. As with any other diffusion process in some space, we are usually interested on the ability of a disease to cover larger shares of the population as time marches on. Detailed analysis of deterministic and stochastic SIR models with spatial components [16] indicates the existence of solutions corresponding to *traveling waves* that propagate the disease among point-like processes. It has also been shown that spatial SIR models can account for the effects of long-distance travel by replacing diffusion operators containing local processes with appropriate integro-differential ones that capture non-local dispersal processes [117].

Agent-based models (ABMs) constitute a family of models where sets of active entities (i.e. agents) interact collectively by following prescribed individual rules intended to portray the emergent dynamics of a real social system [20]. In computational epidemiology, ABMs have been used and comparatively evaluated against SIR models of various kinds. Analysis of the behavior of both classes of models suggests that for many purposes the two classes will give qualitatively the same result, but that agent-based models have an advantage in ease of accounting for heterogeneity in subpopulations where that is significant [5, 96]. Furthermore, the SIR differential equations model is derivable from asymptotic limit of an SIR ABM model through diffusion approximation [17].

A notable coordinated effort to develop agent-based models of a flu pandemic was the Models of Infectious Disease Agents study funded by the National Institutes of Health [51]. More recently the attention of the world was dramatically drawn to the need for public health interventions in the case of COVID-19 by a simulation model projecting 2.2 million deaths from COVID-19 in the United States, and 510,000 in the U.K., in the absence of such interventions [41]. Since then, models continue to be refined as more data are analyzed [3]. It is important to note that there are enormous local geographic variations in the incidence of, and deaths from, COVID-19 [23]. These local variations imply a need for local models, to enable local authorities to construct appropriate strategies of social distancing and testing for mitigation of the effects of COVID-19. The work described in this paper is designed to meet this need.

## 2 COVID-19 poses a wicked policy problem

*Wicked* policy problems are characterized by 1) complexity of elements to be accounted for and their relationship to each other, 2) uncertainty in relationship to the description of the problem and the consequences of actions, and 3) divergence within the affected community of values and interests [55]. By all three measures of wickedness –complexity, uncertainty, and divergence-COVID-19 is a highly wicked problem and will continue to be at least until there is an effective and universally available vaccine.

Dimensions of complexity in COVID-19 emerge from all of the multiple ways in which people interact with each other in such a way as to breathe the same air, and from the consequent trade-offs. These trade-offs involve public health, economics, every aspect of community life, and levels of emotional stress in individuals—a multi-level hierarchy of perspectives involving psychology, sociology, economics, health care, and politics. Correspondingly, we expect COVID-19 modeling efforts to include a growing number of these concerns while remaining actionable and scientifically useful. We expect the complexity of such models to grow, but to do so in a manner that remains intellectually transparent [38] about what is stated in them. To this extent, models become critical components within the top-level decision support system necessary to regain situational control during the current pandemic.

Dimensions of uncertainty abound. As noted above, there is enormous geographic variation in the documented impact of the disease, and variation even in the apparent fundamental parameters of the virus –transmissibility, latency, and virulence-for reasons that are not yet understood. Contributors to the uncertainty may be genetic variation in human populations [26], genetic variation in the virus as it continues to evolve [107], variation in childhood vaccine regimens from one nation to another [82], variations in weather patterns [61], and variations in testing rates and disease reporting accuracy and criteria [60]. Also, there is an element of pure chance –whether or not a particular community was “seeded” with infectious individuals, and how many.

Dimensions of divergence are in some ways clear, and in some ways complicated. The clearest divergence is between the imperative to save lives by social distancing and the costs of social distancing to the community—both economic costs [15, 110] and also costs that are less tangible due to how wealth moves across the global economy [79]. Early on in this crisis most of the world appears to have made the choice of that we would throw our economies into Depression [12] and restrict many community activities we value [40, 97] in order to save the lives of the probably less than 1% of the population who would die from infection should no social distancing constraints be imposed. At the time of writing, this choice is constantly being reconsidered, or at least recalibrated [50]. More, and more open discussions are being held on increasing economic activity even at the acknowledged cost of more infections and even deaths. One is reminded of a famous comedy routine when comedian Jack Benny, whose comic persona was as a notorious cheapskate, is held at gunpoint by a robber who demands “Your money or your life!” This is followed by a long silence, a repeated demand, and a response by Benny, “I’m thinking!”. It seems that COVID-19 has the whole world thinking about the trade-offs between economics –and other aspects of community life- and lives. With respect to divergence, COVID-19 seems as wicked as possible. The economic dimensions of community life can be measured in dollars. The many other dimensions have no common units of measurement, so their relative value is literally incalculable. And yet we are forced to decide about what to value.

Another way to look at a wicked policy problem is a one where the space of potential solution alternatives contains far more social dilemmas than solutions. We may thus define a *social dilemma* is a situation where multiple agents have (explicit or implicit) stakes in the resolution of a problem, stakes are tied to multiple value systems (and not just shared, “objective” technical considerations for example), and a proposed solution contains value contradictions that get translated to unacceptable potential losses were that solution to materialize; at the same time, the social dilemma also provides consequences if a solution fails to appear. Conversely, a perfect solution to a wicked problem is a point of total satisfaction of constraints at all levels of representation of the problem. This is, of course, an idealization; in practice some constraints must be eliminated, relaxed or ignored to find a collective solution. In summary, a social problem is wicked if the density of true solutions is low in the space of all solution alternatives and the search for them can be described as unstructured or even counter-incentivized at best.

Thus, the final element of COVID-19 as a truly wicked problem is that, although it is insoluble, we must make our best effort to solve it. The consequences of not trying to solve this intractable problem, of simply guessing at answers guided only by intuition, are far worse than the consequences of being guided by imperfect models.

## 3 Building a multi-objective model for COVID-19: the agent-based route

Based on the discussion above, our current research efforts have focused on the development of an integrated simulation model capable of a) accurately reflecting known dynamics of the current pandemic and the qualitative results of other models, b) simulating data-driven stochastic heterogeneity across agent populations to more realistically reflect the variability of underlying human populations when the model is applied, c) integrating economic considerations in association with observable features of the pandemic, d) allowing detailed simulation of known public policy measures at different times, intensities and dates, and e) providing a simple interface for non-expert users to configure and interpret.

In relation to the latter point (e), we envision assisting the decision-making process in two steps: first by providing some metaphor or visual proxy for users to construct intuitions by running specific scenarios, and then by translating some of these intuitions into fully-fledged computing and analysis tasks. Succinctly, we wish to facilitate decision making processes that are both robust and adaptive [66] while helping decision makers to avoid falling into the “illusion of control”, or the false belief in the causal relation between computing consequences with a model and immediately improving their decision-making abilities [63]. At the same time, we remain painfully aware of the intrinsic difficulties posed by imperfect data, imperfect implementation of public policy measures, and uncertain timelines for when and for how long to apply measures under unknown timelines for availability of vaccines [104]. Ee expect our model to be beneficial when a) decision makers are fully aware of the underlying simplifications we have made, b) model outcomes are contrasted and adjusted with incoming data during an unfolding situation, c) experts assisting decision makers carefully determine and document how data produced by these simulations is analyzed and translated into tentative recommendations [70].

To this extent, we have focused our efforts on providing modeling tools for population centers with 100,000 inhabitants or less. Our choice is motivated by the geographic distribution of cities and towns across the US [45] and by the apparent inverse correlation between population size and rurality. This is significant since the push for urbanization seems to have driven rural cities and towns to more precarious health systems than their urban counterparts [95]: one can expect COVID-19 propagation to be slower due to lower population densities, but the impact to be at least similar or stronger due to age distribution and availability of health care facilities [8], with a special emphasis on availability of ICUs [58].

### 3.1 Generalities

In our model, agents interact and traverse a discrete 2D torus composed of connected lattice points that represent geographic locations. Agent actions and decisions are governed by random variables with suitable distributions. A single execution (i.e. a *scenario run*) of a parametrization of the model corresponds to a possible world, while a simulation (i.e. a *scenario*) comprises an ensemble of multiple executions with the same parametrizations where outcomes correspond to distributions of agent states and observable quantities must be computed through averages.

The choice of geometry presupposes that agents move across a common landscape at all times, and no agents enter or leave it. This simplification of the geographical landscape, while in general unrealistic, is not uncommon [93, 9] and provides two advantages: a) it naturally matches intuitions behind interactive particle systems driven by mass action principles [75] such as in the SIR model and b) when translated into code, no boundary checks need to be performed by the agents. Lattice sites are connected, from the perspective of an agent, by a Moore neighborhood in an effort to reduce the effect of discretization artifacts [64]. We note here that our model presupposes a homogeneous population density as a means of ensure representativeness of processes within the geographical domain. Although accounting for variable population density areas in the same scenario is possible, our approach simplifies implementation aspects and prevents artifacts for model outcomes that may be strongly density dependent [121] in both epidemiological and economic aspects.

Our model does not explicitly contain a rich representation of locations where agents are drawn to and act as temporal sinks. Instead, we chose to model agent tropism through randomized agent dwell times. *Dwell time* (*τ*_*dw*_) refers here to the minimum amount of time an agent susceptible to COVID-19 contagion needs to spend in a given location to acquire the virus. Based on existing estimates [36], we have chosen *τ*_*dw*_ = 15 minutes, which corresponds to one time step *t*_*s*_, *s* ≥ 0. Average total dwell time 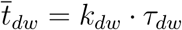 for an agent corresponds to the (integer) average number of steps an agent will dwell on a single location. Since our model assumes a day as a usual reporting unit in decision-making activities, all parameters stated in days are internally rescaled to *τ*_*dw*_ units (i.e. 1 day = 96 simulation steps). Agent dwell times are set at creation time using a random deviate from Poisson(*k*; *λ* = *k*_*dw*_).

### 3.2 Core parameterization

To configure of a scenario, a collection of demographic and disease parameters must be specified. Age structure appears to be strongly associated with differences in COVID-19 fatality rates [19, 32, 37, 98]. The model requires estimates both of the distribution and observed fatality proportions 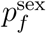 and 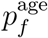 per sex (i.e. male and female) and age (i.e. every ten-year intervals) respectively. Co-morbidities are introduced in a similar manner by means of the age and sex structure of the population as a collection of positive multipliers 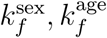 per relevant condition, and aggregate clinical data about their prevalence per age and sex.

The total number of agents *N* at the onset of the simulation remains constant at all times, except when an influx of new agents is simulated. To do so, the number of new agents entering the population correspond to a proportion *p*_new_ of the existing ones. In addition, one must specify when the agents will be introduced *t*_*s*_ = *T*_new_ and how long it takes them to enter the space *τ*_new_. In this manner our model can account for seasonal population increases driven by, for instance, agricultural production or the start of semesters in university towns. After time *t*_*s′*_ = *T*_new_ + *τ*_new_, the simulation will contain approximately *N*′ = *N* + *p*_new_ · *N* agents.

To account for population density *ρ*_pop_, the model specifies the width *W* and height *H* of the lattice that will contain the agents. We presuppose that agents move across the lattice one jump at a time if their dwell time is exhausted. The size of the lattice should be also adjusted based on the mobility and transportation patterns of individuals within the enclosed region of interest –that is, excluding realistic density fluctuations due to commuters that spend most of their time outside of the simulated region. The effect of such adjustment is equal to re-scaling the population density by a mobility pre-factor *k*_mobl_. After these calculations have been performed, the model should ensure for *T*_new_ = *∞* that

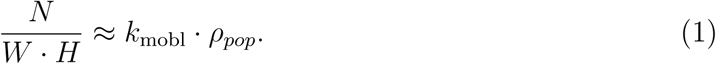

Intuitively, the effect of greater mobility is equivalent to increased population density, or correspondingly, to traversing a smaller space. While our model does not include the effect of road networks or vehicle use, a carefully constructed average for *k*_mobl_ can provide a sufficiently adequate approximation.

Disease-wise, the model comprises six critical parameters. First, the initial proportion of agents *p*_iexp_ that are exposed to the disease. The model assumes that their introduction occurs at the onset of the disease incubation period. Then, the incubation period *τ*_incb_ and the recovery time *τ*_recv_ are inputs correspond to average observed or estimated values in clinical patients [69]. In the simulation, each agents is initialized with individual incubation and recovery times drawn from Poisson(*k*; *λ* = *τ*_incb_) and Poisson(*k*; *λ* = *τ*_recv_) respectively. Our reasoning behind this choice rests on the fact that a) the time at which symptoms manifest across patients depends on a common organismal response to the pathogen dependent on the activation of known (and yet unknown) molecular pathways [2, 103, 111], and at the same time on the intra-population variations that are found across individuals due to their specific genetic make-up and context. Thus, we treat symptoms onset as a homogeneous Poisson process for simplicity even when the described coupling exists. An improvement to our current model would entail, if the reasoning above holds, computing observables using a general Poisson point process by assuming that the Radon-Nikodym density exists [29].

Fourth, the proportion of individuals who remain asymptomatic *p*_asym_ is accounted for and utilized when stochastically deciding the fate of exposed agents. The role of asymptomatic patients remains heavily investigated [11, 85] and appears to play a crucial role in disease mitigation for COVID-19 [18, 33, 43, 122]. From literature data, the proportion of asymptomatic patients appears to vary greatly across countries and demographics (e.g. [4, 6]), although the matter is far from settled. In this sense, our model is intended to be applied by using data starting at the most local level if possible, and only moving to larger geographical instances when data cannot be obtained by means of statistically robust antibody testing.

Fifth, the proportion of severe cases *p*_sev_ is significant for the model due to its relation to hospital capacity. The definition of severity used here is that reported in [118]; we suggest similar guidelines on estimating the proportion severe cases should be followed. Another proxy for severity may comprise the number of non-ICU and ICU admissions, and their ratio [86]; in general, the need of hospitalization implies that clinical evaluation of a patient raises enough concerns as to consider the possibility of transitioning from non-ICU stage to the ICU stage of care [94]. To account for saturation of health care services, the model males use of the proportion of beds proportional to population density *p*_beds_, and we assume that only severe cases are hospitalized. If a severe patient cannot be hospitalized due to saturation, then its probability of fatality rises by a factor to be determined empirically; for reference, our model presupposes a four-fold increase. At present, our model does not provide estimates of ICU occupancy.

Finally, the model utilizes the probability of contagion *P*_cont_ per agent per interaction. This quantity can be obtained from field data or other (more coarsed-grained) epidemiological models at the onset from estimates of the basic reproduction number *R*_0_ by observing that

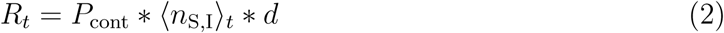

where ⟨*n*_S,I_⟩_*t*_ is the average number of contacts between susceptible (*S*) and infectious (*I*) agents, and *d* is the duration of infectiousness of the disease. Recalling the SIR model

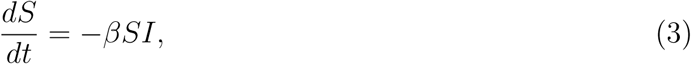

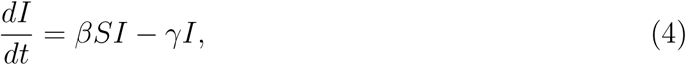

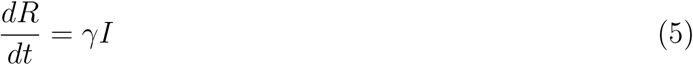

we observe that *γ* = *d*^*−*1^ and

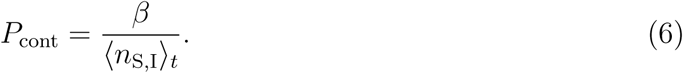

Our view of *R*_0_ is that of a preliminary estimate for initial calibration at the onset of the period of interest. For reporting purposes, we favor the effective reproductive number *R*(*t*), and consequently provide information about the observable for *n*_S,I_(*t*) such that

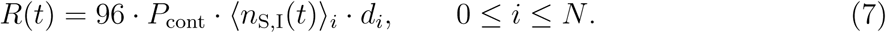

Since one agent represents multiple individuals in the region of interest, it becomes necessary to compensate for this renormalization process. For such purpose, we provide a scaling factor associated to the representativeness of the model *k*_*r*_ given with a scale 1:*R* by

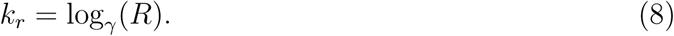

We found empirically *γ ≈* 1.58489, such that rescaling the probability of contagion to obtain 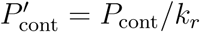 leads to

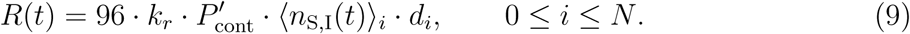

In addition, *R*(*t*) ∝ *ρ*_pop_ (Eq. 1), which implies that *k*_mob_ must also modulate *R*(*t*). The final expression becomes

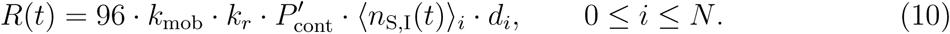

### 3.3 Agent dynamics

At the model level, observables are interrogated across the agent population, agent step actions scheduled and the step number updated. At each step, agents move one space across the 2D torus after exhausting their dwell time per location. All agents posses an internal state *σ* that stores information relevant to the disease, its economic aspects and various control structures. Their motion is driven by a random walk within their Moore neighborhood, unless their state has been set to isolation. Isolation means not moving across the space regardless of dwell times. More than one agent can inhabit one lattice site, which forms the basis for determining when a susceptible agent becomes exposed and the infectious cycle starts. Prior to performing stage-dependent computations from the disease perspective, agents compute the consequences of policy measures and adjust various elements of their internal states relevant to epidemiological and economic actions to follow.

#### 3.3.1 Epidemiology

Our model departs from the usual compartments of the SIR and extends it in order to account for a more fine grained variety of significant infection stages. Agent disease states are as follows:

##### Susceptible

All agents (except those marked as initially exposed) start as the susceptible population. When susceptible agents share the same lattice site, these may come in contact with other exposed, symptomatic (i.e. due to voluntary or involuntary breaking of quarantine with probability 1 − *P*_isoeff_) or undetected asymptomatic agents. If at least one agent is infectious, the agent changes its state *σ* to exposed as dictated by Bernoulli(*σ, P*_cont_). Unless quarantined due to a policy measure, susceptible agents move freely across the lattice.

##### Exposed

In the absence of any policy measures impacting exposed agents, these continue to explore lattice sites until their incubation period given by Poisson(*x, λ* = *τ*_incb_) is exhausted. At that time, agents become asymptomatic as dictated by Bernoulli(*σ, p*_asym_) or symptomatic detected, otherwise.

##### Asymptomatic

Asymptomatic agents continue moving through the space and remain infectious until the recovery period given by Poisson(*x, λ* = *τ*_recv_) is exhausted. At that point, the agent enters the population of those recovered.

##### Symptomatic

When an agent becomes symptomatic, it is immediately marked as detected and quarantined. Regardless of stringency of testing policies, the definition of confirmed case depends at the minimum on being both symptomatic and a positive identification via some form of testing (i.e. RT-PCR qualitative or quantitative, serological [91]). Symptomatic agents follow two possible trajectories. In the first one, agents convalesce without becoming severe until their recovery time is exhausted an recuperate. In the second one, agents become severe as dictated by Bernoulli(*σ, p*_sev_). In terms of the impact of saturation of health care services, research is needed to determine lethality of patients outside hospitals and other facilities. However, using existing ethical guidelines that provide heuristics of fair resource allocation for beds and ventilator equipment [39] as a proxy, we estimate that lethality increases (conservatively) by at least a factor of 5.

##### Asymptomatic Detected

Asymptomatic agents detected by some widespread systematic testing strategy are immediately quarantined in place, and wait until their recovery time is exhausted. At that point, they join the population of recovered agents.

##### Severe

Agents that enter the severe stage represent patients that require some form of hospitalization, and for some of them, use of mechanical ventilators; these remain under perfect quarantine. Lethality, computed per age and sex population structures as 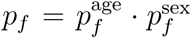 is used to determine whether a severe agent becomes a fatality as given by Bernoulli(*σ, p*_*f*_). Agents, on the other hand, may survive until recovery.

##### Recovered

Agents that have recovered leave quarantine and move again freely across the lattice. Our model does not consider the probability of re-infection, but this may need to be included in the future [92]. At the moment of writing, this aspect of COVID-19 remains speculative and uncertain for human patients [34, 90, 115] despite encouraging evidence obtained from experiments using rhesus macaques [14].

##### Deceased

Agent that count as fatalities do not interact with other agents or undergo any further epidemiological significant events until the end of the simulation.

#### 3.3.2 Inbound infectious cases

In order to increase model realism, we consider the effect of inbound infectious cases of two sorts: people that live within the community but have to travel and work outside of it in a steady stream daily that maintains the overall population density stable, and people that move seasonally within the community, potentially bringing in more cases that have a different viral load. This last case describes, for example, the opening of university campuses for instruction where several people relocate to adjacent towns.

For the first situation, the model includes the probability of susceptible people becoming infected with a daily rate that determines the infectious stage depending on Bernoulli(*x, r*_ibnd_). In the second type of infectious cases, at time *T*_mass_ a proportion *p*_mass_ of the current population will enter the simulation space during a time period *τ*_mass_ with a probability of being in the exposed state of *P*_mass_.

### 3.4 Policy measures

COVID-19 has forced a frantic search for public policy measure combinations capable of containing viral transmission, and ideally quelling its progression altogether [13, 42, 47, 56, 67, 71, 89, 99, 106, 115, 116, 120]. All measures reviewed and emerging across literature roughly belong to four main classes of measures: 1) those that aim to reduce at any instant the density of individuals at locations with potentially high concentration of people by means of imposed self-isolation of non-essential workers and cancellation of activities involving massive amounts of people, 2) those that reduce the likelihood of viral exposure for those qualified as essential workers for whom close social interactions are inescapable, 3) those intended to detect and isolate positive virus carriers through application of molecular or serological testing and 4) those that seek to reconstruct interaction histories in which a positive infectious patient may have had an active role in unknowingly spreading the disease.

It has become increasingly clear that these measures are essential yet hard to sustain for long periods of time. On the one hand, various degrees of negative psychological impact have been recently reported [31], in particular impacts that decrease adherence to public policy measures [59] which are expected to naturally arise after periods of prolonged confinement under a collective crisis; World War II critics of air raid shelter policies constitute a significant precedent [80]. On the hand, mounting concerns on lasting economic impacts [76, 83] materialize the wickedness of the COVID-19 pandemic and the cost of measures to address it [22], concerns that which include social protection of workers [44], the labor market [30], patterns of work [65], gender equality [7], nation-state economics [74], and monetary policy [21] to name a few.

Motivated by the latter, our work attempts to model the individual variability expected when these types of measures are implemented, their various impacts in terms of disease and economy collective observables, and the potential outcomes of combining them in various manners. We note that societal and economic impacts of COVID-19 differ from those in other pandemics due to the tight coupling of global events and the effect of near-instantaneous digital communication. *We are changing the pandemic while living it*. While our model does not provide mechanisms to state the associated control problem in cybernetics terms, emerging literature (e.g. [35, 48, 87, 119]) suggests that such approach may be possible, and even essential to provide solutions that account for the complexities involved in politically and socially driven environments.

#### 3.4.1 Self-isolation

Self-isolation in our model is captured by establishing a period in which a proportion *p*_isol_ of agents in mobile states (i.e. susceptible, exposed, asymptomatic) remains at a fixed location for a well-defined period of time. Self-isolation starts at time *T*_isol_ and extends for a period *τ*_isol_, after which motion across space is restored. Agents isolate with effectiveness *P*_isoeff_, representing the probability that when in contact with another infective agent the final probability of contagion becomes 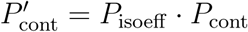.

#### 3.4.2 Social distancing

Social distancing is modeled as a distance-dependent adjustment constant *δ*(*ℓ*) that adjusts the probability of contagion *P*_cont_ depending on linear distance *ℓ* assumed between agents within the same cell such that 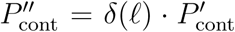. Based on recent experiments on the effect of air turbulence on droplet dispersion [24], we assume a decreasing sigmoid profile after 1.5 meters. Hence,

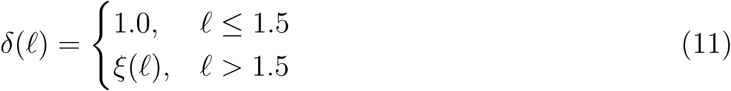

with

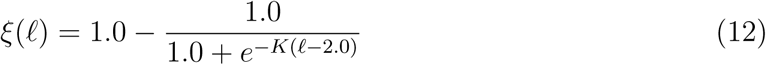

where *K* is a constant that adjust the decrease rate of the sigmoid function. For the purposes of the COVID-19 model, *K* = 10.0. The value of *ℓ* can be adjusted also to account for the effect of other interventions that decrease contagion probability per contact by decreasing the effective viral load, such as the use of various types of face masks [62].

#### 3.4.3 Testing

Similar to self-isolation, testing in the simulation operates by distributing a target percentage of the population into a given period. Susceptible and asymptomatic agents can be tested; in our simulation, we do not re-test those who recover. A symptomatic case is treated immediately as tested, representing the case where a patient reaches a health provider and a test is applied to determine the correspondence between symptoms and the disease.

Testing proceeds in the following manner. Once time *T*_test_ is reached, symptomatic and asymptomatic agents are selected with a probability *P*_test_ proportional to the period *τ*_test_. This testing process simulates massive testing policies without any statistical design or underlying population structure.

#### 3.4.4 Contact tracing

We simulate forms of automated contact tracing by means of a set of known prior contacts. We assume a delay of two days for contact follow-up once an infected patient has been discovered. Once an agent is marked as positive, all of its contacts are evaluated and classified either as susceptible (negative), symptomatic or asymptomatic detected. Contact tracing utilizes the same start time and period as testing.

### 3.5 Estimation of economic impact

Along with an epidemiological model, we have included economic factors tied to disease stages. After some investigation around statistical theories in economics [27], we decided to implement a simple model where value creation in terms of exchanges between money and products or services [84] are computed in connection with the progression of the disease. Our model is an oversimplification of economic systems, and as such, its goal is to provide a numerical intuition about the immediate effects on the accumulation of capital by individuals and the public sector during the period of interest. Thus, the economic model makes no assumptions about wealth distribution, wealth inequality or other societal factors, and as such only aims at portraying the impact of an epidemic on transactional capital gains or losses at the private and public levels.

Regarding public value [81], we observe that the complex web of actions across individuals and institutions make construction of a detailed model expensive in connection with infectious disease dynamics. Growing literature on the subject is indicative of the latter [25, 53, 78, 112]. To the best of our current experience, proper treatment of the current situation would require a different class of model based on fractional operators coupling epidemiological [1] and econometric aspects [109, 73] capable of accounting for short- and long-term memory macroeconomic effects [108].

#### 3.5.1 A disease-economy input-output system

Our take on the matter can be stated through the following somewhat intuitive principles:

1. Disease stages that allow interactions should cause non-linear positive externalities in terms of collectively amplified public value. The more frequent interaction exchanges, the higher the materialization of public value as a function of non-linear effects of reaching throughput efficiencies that both maximize economies of scale and translate into deep capital accumulation and redistribution across the public sector.
2. Disease stages that forbid interactions but do not put individual lives at risk should cause linear negative externalities associated to both increased unemployment ripple effects and inability to reach throughput thresholds capable of amplifying value creation.
3. Disease stages that both forbid interactions and put individual lives at risk should cause non-linear negative externalities due to increased unemployment ripple effects, inability to reach throughput thresholds capable of amplifying value creation and the saturation of alternatives under an increasingly severe public health crisis.
4. In addition, we estimate the cost of performing one test as part of the public cost. The purpose of this observable is to provide an account of testing as a public measure versus other actions for which their cost is harder to account for.

In order to materialize the above principles, our approach is inspired by that of input-output matrices utilized to account for combined economy-ecosystem calculations [52]. The equivalent of the matrix ***M*** in our model expresses economic outputs per disease stages. Viewed as a matrix computation, the components of the input vector ***u*** correspond to proportions of the agent population per infectious stage, while the output vector ***υ*** is of the form ***υ*** = (*υ*_priv_, *υ*_pub_). Depending on the disease stage, vector components are computed aggregating the value of interactions or individually. By a suitable homotopic transformation 𝒯 [88], we approximate non-linear effects of market dynamics, thus the final value of ***υ*** becomes 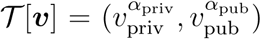. The matrix components ***m_ij_*** and both *α*_priv_ and *α*_pub_ are model inputs.

### 3.6 Model outcomes

Our model captures during its execution various observables every time step. All values are aggregates and no particular agent information is stored; in the future, this can be of value if the model is extended with network information. After a simulation has completed, the following classes of observables are recorded in a CSV format:

**Table 1:** Example of a coupled disease-economy input-ouput matrix with *α*_priv_ = *α*_pub_ = 1.

| Disease stage | Private value | Public value | Computation |
| --- | --- | --- | --- |
| Susceptible | 1.0 | 1.0 | Aggregate |
| Exposed | 1.0 | 10.0 | Aggregate |
| Asymptomatic | 1.0 | 10.0 | Aggregate |
| Symptomatic | -0.2 | -3.0 | Individual |
| Asymp. detected | -0.2 | -1.0 | Individual |
| Severe | -5.0 | -15.0 | Individual |
| Recovered | 0.8 | 2.0 | Individual |
| Deceased | 0.0 | -0.2 | Individual |

#### Simulation

Step number, population size

#### Disease (fraction)

Susceptible, exposed, asymptomatic, symptomatic quarantined, asymptomatic quarantined, severe, recovered, diseased

#### Measures

Self-Isolated, tested, traced

#### Epidemiology

Effective reproductive number

#### Economics

Cumulative private value, cumulative public value

### 3.7 Implementation

The Epidemiology Workbench is implemented using Python (v 3.6) using the Mesa agent-based simulation library [77]. In batch processing mode, our model receives a JSON file with all the parameters described above and a number indicating the number of cores to use during the simulation. Once a sanity check is performed, the parameters are used in conjunction with the multiprocessing batch running features provided by Mesa.

We also provide a parameterizable web-based dashboard to explore individual runs. The objective behind this corresponds to enabling decision-making users to progressively gain intuitions behind each parameter, and not to provide operational information. Our code is openly accessible through GitHub^1^.

### 3.8 Calibration

Model calibration requires the best possible clinical data estimates for three critical parameters: the average incubation time, the average recovery time, and the proportion of asymptomatic patients. For the average incubation time, hospital triaging of COVID-19 cases can provide information leading to estimates, or values provided by trusted sources at the most local level possible can be used. Incubation time is likely to be blurred by multiple confusion variables captured in our model as a Poisson distribution. Despite its stochasticity, the model supposes that large population sizes group tightly towards a mean value. This assumption may need to be revised retrospectively later. For the proportion of asymptomatic patients, contact tracing and structured antibody testing can provide specific local information.

As a general calibration protocol, we suggest the following steps:

1. Adjust grid size based on fixed population density until *R*_0_ matches the best known value for the area in the first days of the model (steps = 96) and *>* 30 runs. Population density in the model loosely includes average mobility patterns, and cell sizes reflect the distance traversed every 15 minutes. Also –but not recommended- the probability of contagion may be used to calibrate. This may imply unknown population conditions and should be used only to test hypotheses about individual variations that manifest in the ability of COVID-19 to spread.
2. Execute the model to the point where the number of symptomatic agents corresponds to one representative agent. For example, with an ABM of 1000 agents and a population of 100k individuals, the critical infected agent-to-population ratio is 1:100. Use the point in time rounded to the nearest integer day as point of departure for policy measures. In practice, this implies executing the model in excess of as many days as the longest known incubation period.
3. If policy measures have been introduced, use date above as the reference point for their introduction.

To develop scenarios, we strongly recommend starting from the most recently calibrated model that includes policies as well as using a model without measures as a basis for counterfactual arguments.

## 4 Case study: the cities of Urbana and Champaign after reopening UIUC, Fall 2020

The cities of Urbana (pop. 42,214)^2^ and Champaign (pop. 88,909)^3^ comprise a population of approximately 132,000 inhabitants. Figure 1 describes the current percent distribution per age group. In addition, distribution per sex is 50.5% males and 49.5% females. However, the population across both cities undergoes a seasonal decrease on mid May due to the end of the Spring semester and an increase in early August with the start of the Fall semester in the University of Illinois at Urbana-Champaign. From the more than 50,000 students enrolled on campus on May^4^, we estimate that around 30,000 leave during summer to return for the fall semester. Effectively, the summer population amounts to around 100,000 inhabitants. We used COVID-19 age- and sex-dependent mortality values as reported by CDC until June 10.

**Figure 1:**
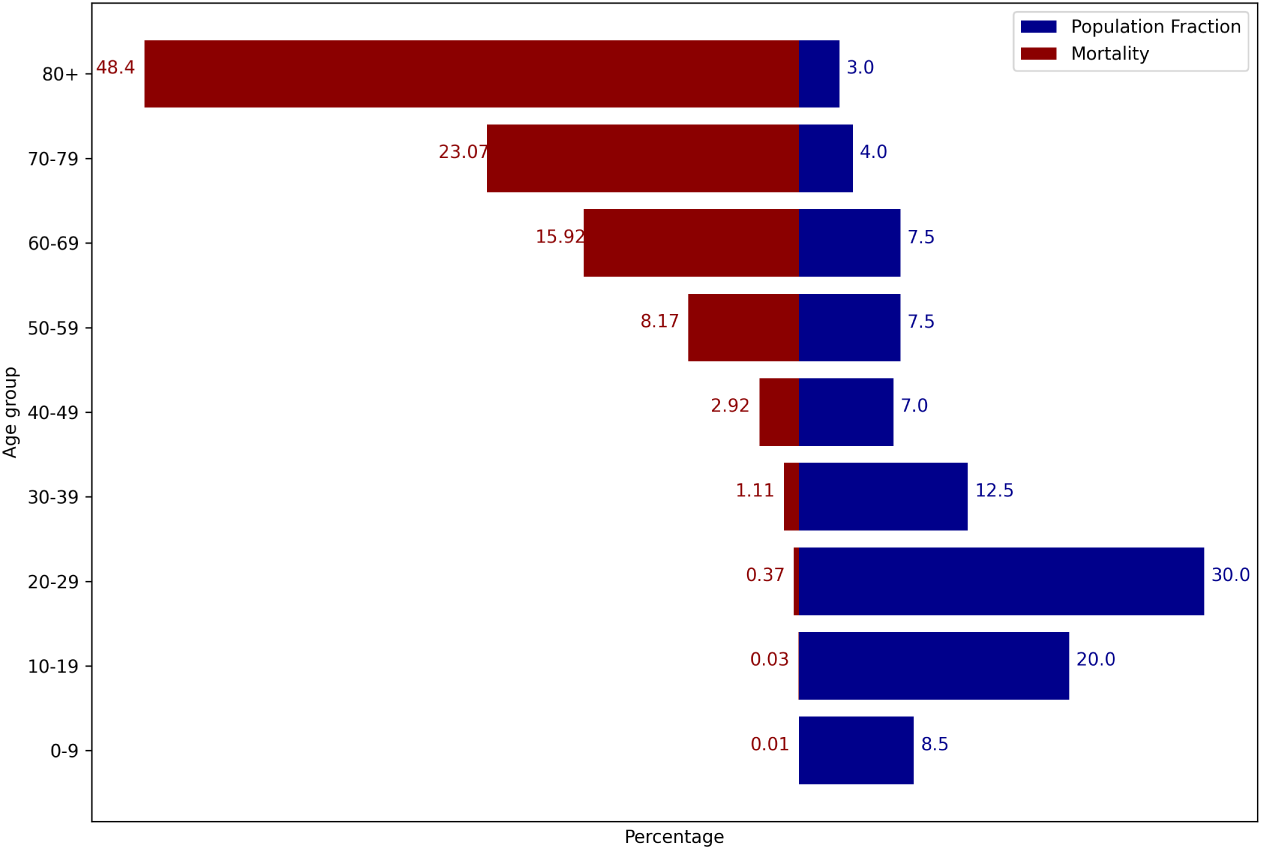
Combined population age structure and associated mortality in Urbana and Champaign cities, 2019 estimate by US Census Bureau/CDC.

Our interest rests on simulating the effect of various measures once mobility restrictions remain at values similar to the present one (phase 4 reopening) and combinations of testing and automated contact tracing when an estimate of 30,000 incoming students with a higher average viral load reach both cities, on an increase of 30% of the population present in summer.

### 4.1 Calibration

COVID-19 data was obtained from the Champaign-Urbana Public Health District (CU- PHD)^5^ and CDC for mortality data^6^. Sex-dependent mortality was established at 61.8% for males and 38.2% for females. The first local case in the community was reported on March 8, and state-wide shelter-in-place measures were applied on March 21^7^. Later on, mandatory mask usage was established on May 1^8^. By April 21, cumulative cases had reached 0.1% of the population, and by July 8 it had increased to 1%. The local *R*(*t*) value at the peak period between April 21 and May 17 reached an estimated value of 1.2; after these measures, it remained around 0.91. We used a probability of contagion per interaction of 0.004 every 15 minutes if there is at least one person infected at the same location as the susceptible one. This value, although computed from data, appears to reflect compliance with various sanitation practices among the population. Based on the fluctuation in local data, we have estimated an inbound probability of 0.0002 new cases due to members of the community becoming exposed elsewhere. CU-PHD performs strict contact tracing across all cases, hence for all simulations we assume contact tracing remains active across all simulations.

Google Mobility data were used to estimate the average effective shelter-in-place value to a 45% of the population with 0.8% efficacy of self-isolation. The severity was similarly estimated at 5% based on local case information. Similarly, a starting value of *R*(*t*) = 1.2 corresponded to a grid of 190 × 225 cells and 1000 agents and *k*_mob_ ≈ 0.4781; this last value appears to be consistent with the regular use of public transport in the area and local observed shelter-at-home patterns. At the start of the simulation, the agent-to-inhabitant ratio is approximately 1:100. Hence, the calibration starts with one exposed agent. Differences between *R*(*t*) in April and July correspond to *ℓ* = 1.89. Intuitively, this implies that the effect of wearing a mask may roughly translate into increasing the distance 0.39 meters beyond what WHO recommends for proper social distancing. However, this cannot and should not be interpreted as to relax mask usage in any manner. In regard to asymptomatic patients, we have established a conservative value of 35% based on prior studies [49]. The following sequence of events was assumed toward the start of classes this Fall:

1. Exposure of the first representative agent on April 15 (simulation day 0)
2. First symptomatic representative agent on April 21 (simulation day 6)
3. Mask order from the State of Illinois on May 1 (simulation day 16)
4. Students start massive ingress two weeks between August 9-22 (simulation days 116- 129)
5. Community members are tested between August 23-29 (simulation days 130-136)
6. Simulation continues without testing for two weeks between August 30-September 12 (simulation days 137-153)

### 4.2 Simulation targets

Our goal for simulating the impact of mass ingress was to determine, based on various measures, the viability of preserving public health under variations of measures a week after a significant portion of the population has been tested. To determine the latter, we obtained data for *R*(*t*), symptomatic, asymptomatic, severe and deceased fractions of the population. In addition, we collected information about economic impact using our input-output matrix model. A total of 6 simulations explore the following parameter settings:

1. shelter-in-place continued/removed on day 117,
2. massive testing performed to 25%, 50% and 75% of the population in the enlarged community.

We also computed a counterfactual case corresponding to no massive testing and lifting of shelter-at-home orders to compare against a backdrop without measures. Model calibration results (Figure 2) correspond to the parametrization publicly available at the GitHub project repository^9^.

**Figure 2:**
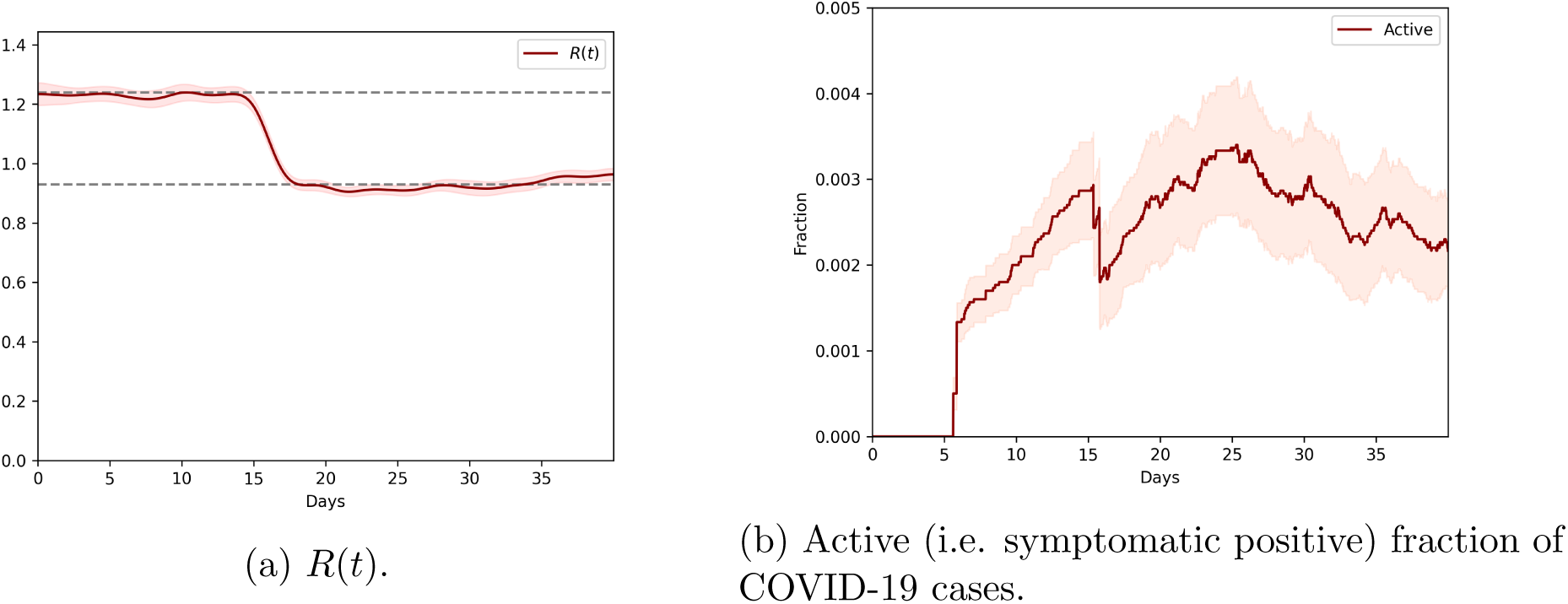
Calibration outcomes for the cities of Urbana and Champaign during the COVID- 19 pandemic. Dates span from April 21 to May 31 (40 days).

#### 4.3 Results and Discussion

We computed a total of 7 scenarios (shelter-at-home times testing levels plus counterfactual), each one with an ensemble of 30 independent runs. Execution of these scenarios was performed on Amazon EC2 infrastructure using a c5a.8xlarge non-dedicated instance with 36 processors and 64 GB RAM. We used an Ubnutu 18.04 86 image as the choice of operating system. Calibration CPU time with an ensemble of similar size for the first 40 days is, on average, 16.3±0.4 minutes, and the average execution time of a complete scenario is 65.2 ± 1.3 minutes. Preliminary profiling indicates that random number generation using the SciPy library [114] explains most of the execution time. No attempts were made to further speed up our code by compiling it using Cython [102] or Numba [68]. Our goal in these simulations was not to reproduce exactly the case curves observed in the community, but to obtain a picture that remains quantitatively and qualitatively rigorous. Confidence intervals are calculated at 95% when present. We provide scripts to automate the setup of the Amazon EC2 instance with model installation^10^, the execution of all scenarios^11^ and their visualization^12^ for reproducibility purposes.

The following convention is applied to all figures: dark red corresponds to 25% testing, teal to 50% and dark blue to 75%; the counterfactual case is colored in purple. A dotted line corresponds to the start of mass ingress, a dot-dash line to the end of mass ingress and the start of massive testing, and a dashed line to the end of massive testing. All figures related to disease stages start at day 80.

#### 4.3.1 Public health

Model results indicate that outbound exposed individuals coupled with local fluctuations appear to drive the behavior of this pandemic for the cities of Urbana and Champaign. The combination of contact tracing and public health management by CU-PHD, compliance with health and sanitary measures and rapid implementation of shelter-in-place measures have prevented the pandemic from escalating in the region. Considering both cities as a closed system, adequate health management appears to be ultimately responsible for the small number of severe cases and hospitalizations in the region. The effect of masks, based on the information obtained from our simulation, appears to have a significant effect on the value *R*(*t*) according to Fig. 2.

Our simulation of the reopening of the University of Illinois local campus with a higher viral load suggests that an increase in cases should be observable independent of the testing regime or relaxation measures. The future impact of this increase, however, is not. Figure 3 indicates that lifting all shelter-at-home restrictions and performing testing has a significant impact regardless of the testing level, while preserving shelter-at-home measures along with any testing level can drastically sustain *R*(*t*) slightly below pre-mask order values. We note that *R*(*t*) decreases in all cases, which can be explained by contact tracing-based testing. This suggests that even hybrid education modes (in presence + online) constitute significantly better alternatives than full campus reopening.

**Figure 3:**
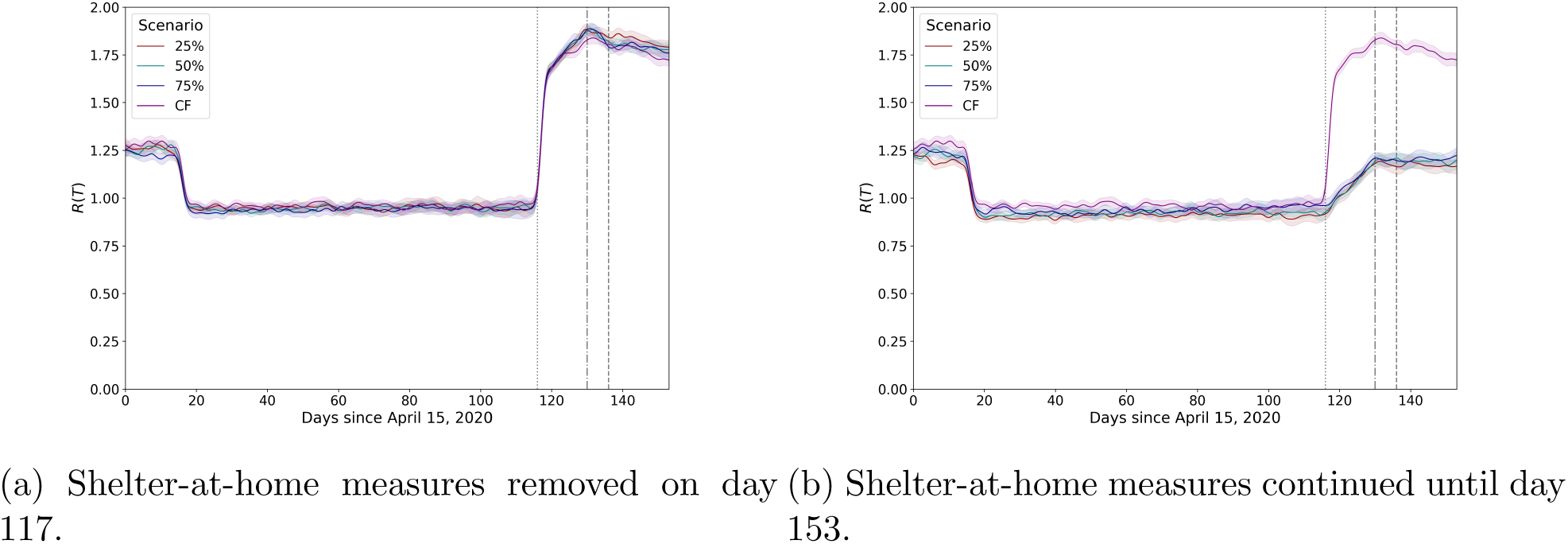
Evolution of *R*(*t*) as a function of testing levels and shelter-at-home measures removal (a) and (b) preservation.

Testing intensity matters. In terms of the outcome after day 137, testing intensity determines the last value of active cases during two weeks after testing. Note that even when the testing instant itself has passed, capturing a larger and larger number of positive cases (particularly asymptomatic ones) drastically reduces the infectious population (Figure 4). Even when shelter-at-home measures have been lifted, testing reduces the fraction of the population classified as active cases at least higher but close to its value prior to mass ingress (25%), roughly equal its prior value (50%) or lower than the prior value (75%). Lifting shelter-at-home measures has a significant impact on the magnitude of both the peak of active cases and the value after two weeks have passed since testing occurred. Plans to test the student population once per week at scale, although not simulated here, appear to be a most effective solution to further tame the curve.

**Figure 4:**
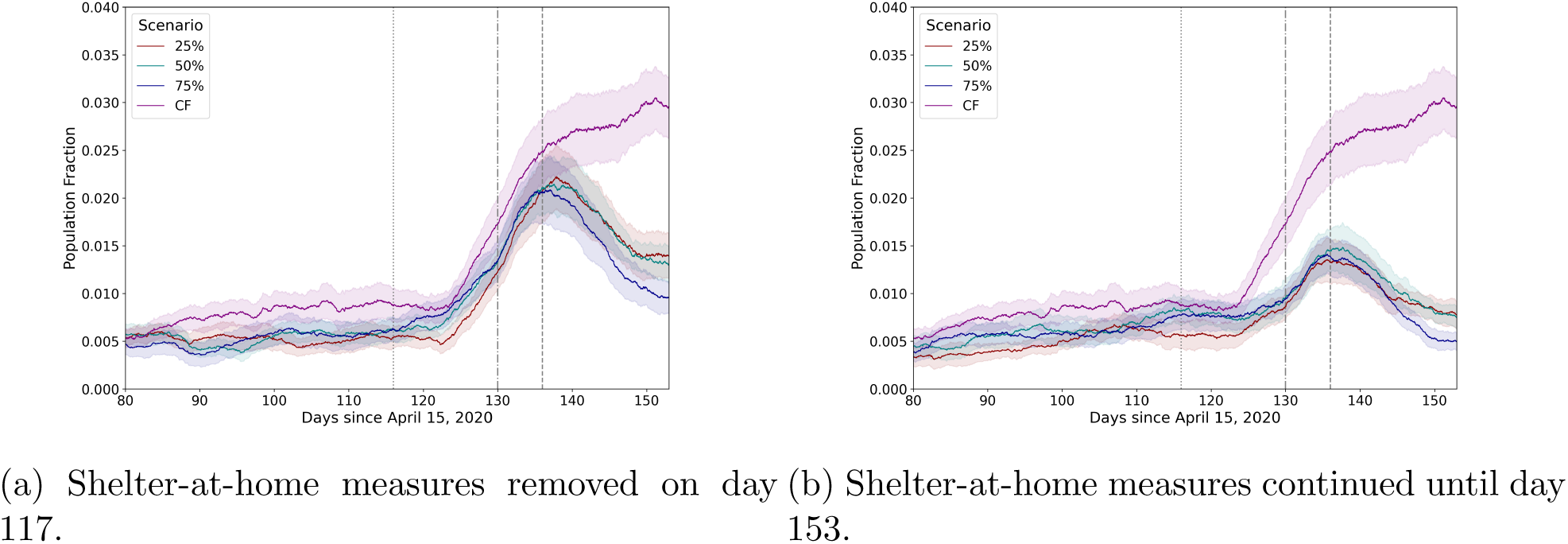
Active cases as a function of testing levels and shelter-at-home measures removal (a) and (b) preservation.

The main mechanism massive testing addresses in general, according to our simulations, is the removal of infectious individuals from the population, in particular those who are asymptomatic. In general, the estimated proportion of asymptomatic patients is a significant driver of the contagion in our model. When the population increases, many more individual contacts are possible within the same geographical area, and the lag induced by the incubation time translates into observing the impact of testing at least a week later. Figure 5 compares the asymptomatic fraction of the population across a baseline simulation without massive testing or some degree of sheltering. As in the previous case, shelter-in-place measures have an effect on the growth rate of the asymptomatic population, but even a testing intensity of 25% appears to lower it significantly compared to the counterfactual case. At case severity of 5%, more hospitalizations may be expected six days after day 130 (mass ingress), particularly if shelter-at-home orders have been lifted (Figure 6). The impact of testing, while it cannot be fully distinguished due to the overlap of confidence intervals in our simulations

**Figure 5:**
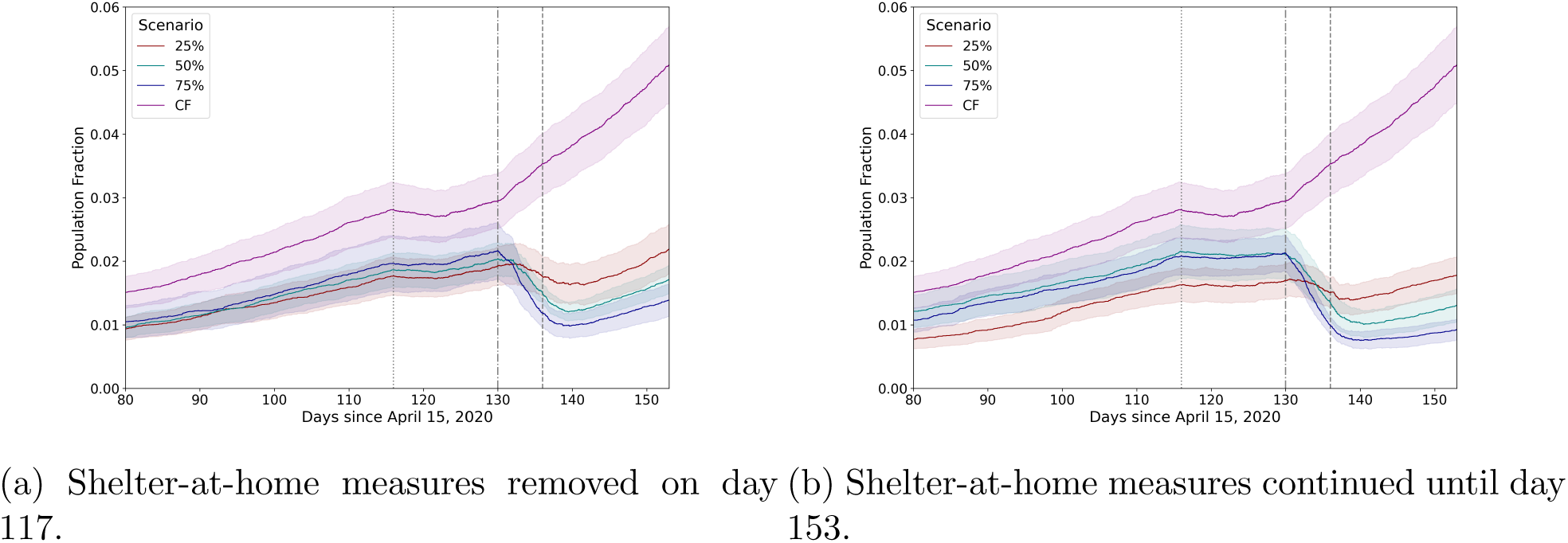
Asymptomatic cases as a function of testing levels and shelter-at-home measures removal (a) and (b) preservation.

**Figure 6:**
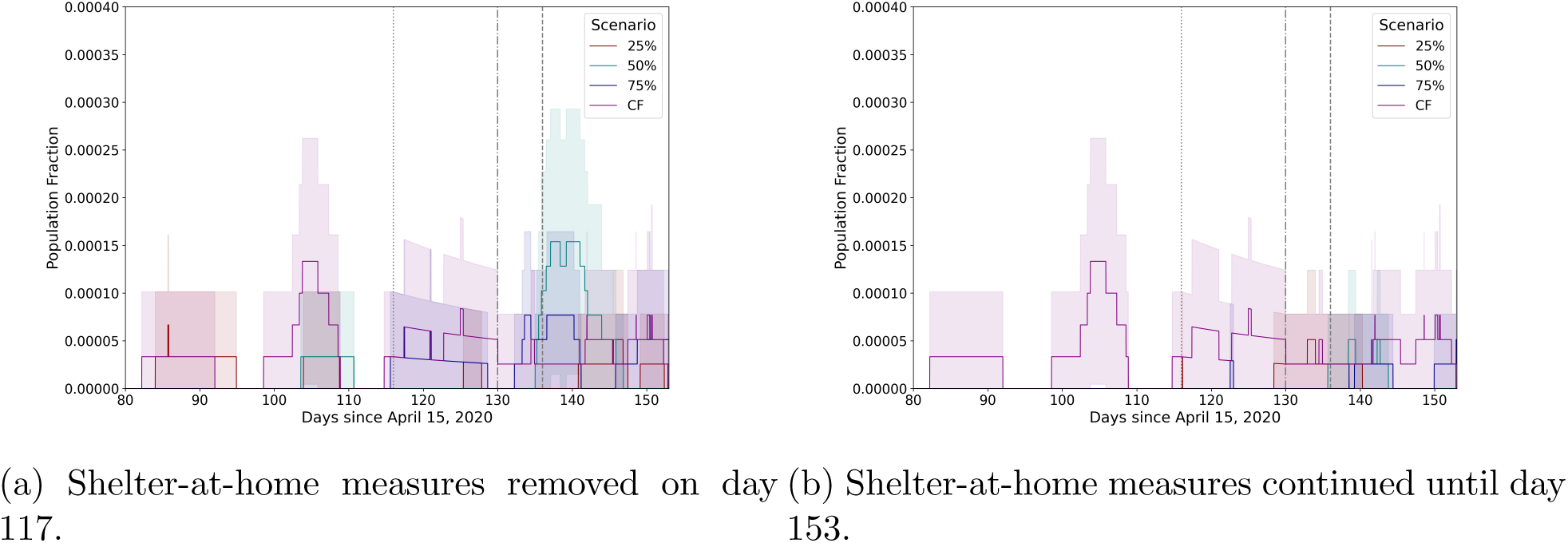
Severe cases as a function of testing levels and shelter-at-home measures removal (a) and (b) preservation.

#### 4.3.2 Economic impact

Our analysis of economic impact focuses on two per-capita averages: cumulative private value (without any egress) and cumulative public value. While this part of our proposed model is experimental and requires further analysis, we proceed to state the current results.

First, we studied the effect of the pandemic only on individual accumulation of private value (Figure 7). Mass ingress appears to renormalize temporarily the distribution and removing shelter-at-home measures, predictably, increases the final value at day 153, but only by approximately five units. All testing levels appear to comprise a relatively tight bundle, which can be interpreted either as an artifact due to the model being simplistic, or as the fact that the impact of testing levels on cumulative private value in the context of a pandemic under control (as in Urbana and Champaign cities) is limited. Even when these differences are small, they do exist. Testing at low levels (i.e. 25% and 50%) reduces the number of isolated people compared to testing at a broader scale (i.e. 70%). However, when an epidemic process remains under control, public health benefits largely appears overshadow individual losses, contingent on the validity of this approach.

**Figure 7:**
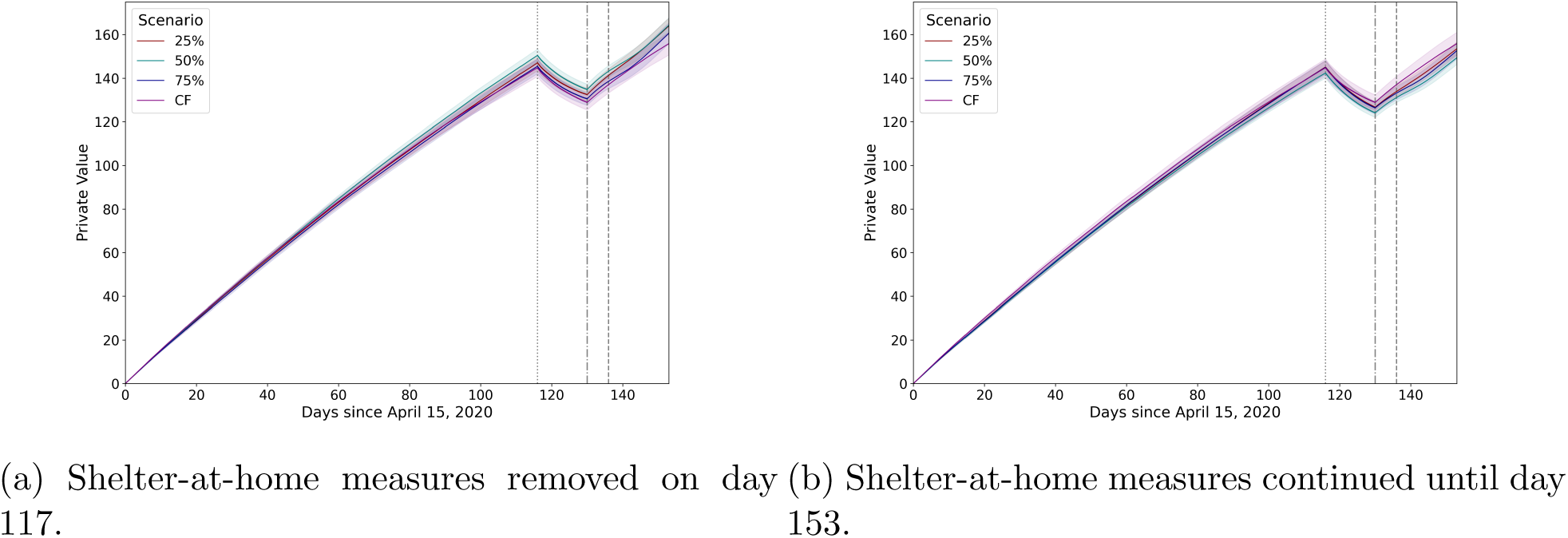
Per-capita cumulative private value as a function of testing levels and shelter-at-home measures removal (a) and (b) preservation.

In the case of per-capita cumulative public value, the model predicts negative outcomes even for an epidemic under control (Figure). This appears to align with public expenditure needed to mitigate any economic and societal lockdown necessary to stop the spread of the disease, as well as the negative externalities leading to less public transactions taking place on systems that were designed to support a certain minimum load to remain profitable: public finances tend to be, in general, inelastic for that reason. When shelter-at-home measures are lifted, a natural order of solutions arises from worst-case (our counterfactual, purple) to best-case (75% testing, blue): strong testing reduces the long-term impact of active, asymptomatic and severe cases. In this situation, however, the effect of mass ingress appears to be to re-bundle the behavior as *R*(*t*) increases again. If shelter-at-home is preserved, an interesting situation arises: doing nothing appears to be a good economic solution. We believe the main reason behind this result to be that, for the case modeled here, the social and economic cost of the lockdown is higher due to the situation being under control; however, the material gains computed by the model between are rather small, and preserving public health over economics is a better-long term strategy. Despite this, strong massive testing still provides a next best solution from the point of view of economics, and the best strategy from the public health perspective; this is evidenced by the diverging curves in Fig. 8(b). We speculate, based on our current simulation outcomes, that the ordering in the economic cost profile of a pandemic during its exponential phase should be similar to that of Fig. 8(a), but with the divergence observed in (b).

**Figure 8:**
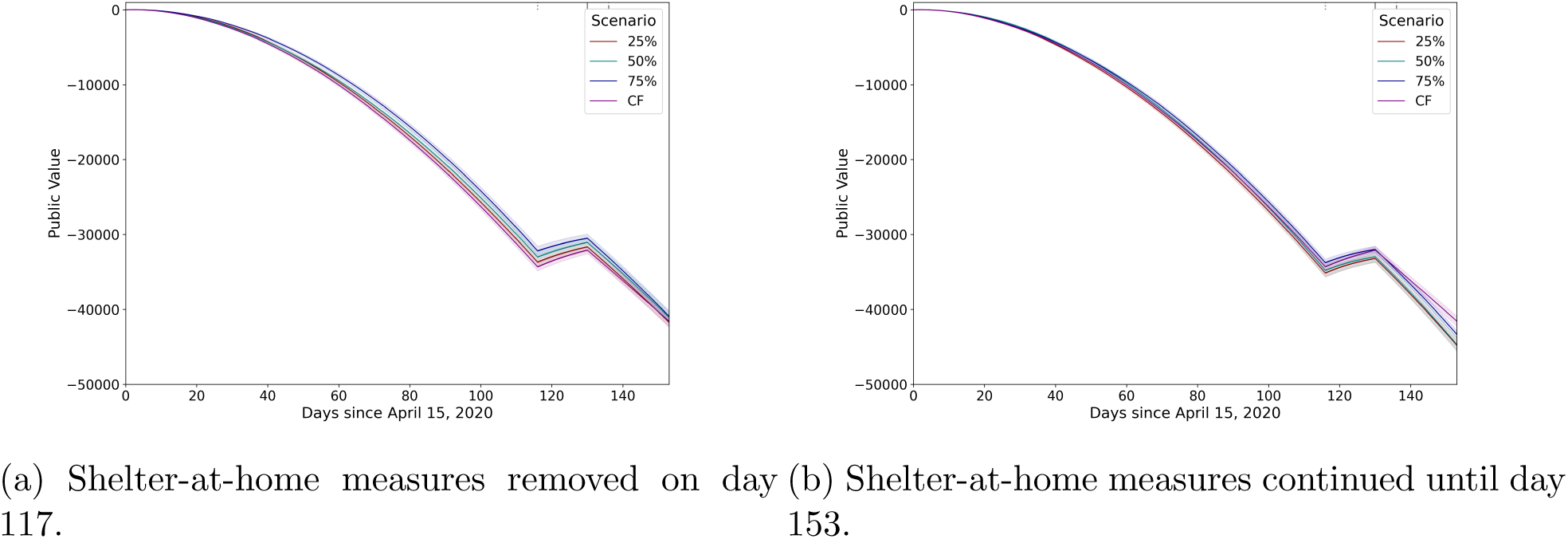
Per-capita cumulative public value as a function of testing levels and shelter-at-home measures removal (a) and (b) preservation.

## 5 Conclusions

We reported here the construction of an agent-based workbench using the Mesa modeling framework capable of capturing epidemic processes alongside public policy measures. The model is fully stochastic, entailing the computation of observables of different kinds. While computationally expensive, its formulation allows to easily obtain quantities that appear to be useful in the process of combating an epidemic. We applied our workbench to understanding the possible epidemiological profile of two cities, Urbana and Champaign, in the context of the reopening of the University of Illinois local campus next Fall. Our simulations indicate that at least 50% testing of the local population is needed to sustain the pressure of mass ingress of individuals with a higher viral load compared to the local one. More generally, contemporary management of an epidemic demands changing the *mode of interaction* across as much as the population as possible from those requiring physical proximity to those that do not. Although digital technologies provide mechanisms to preserve safe spatial distancing, temporal distancing can also be intelligently used to reduce the probability of contagion. In terms of economics, public health measures must be privileged over financial concerns, since the panorama appears similarly bleak during the early phases of an epidemic, and strict measures possibly provide the best solution during the exponential phase.

Our model contains the following key limitations. First, only one measure per type can be specified at the moment, instead of a sequence of dates paired with values corresponding to the measured (or expected) effect of measures of the same type. In the example above, we used an approximation of shelter-in-place for the entire simulation period (April 21– September 12), even when the State of Illinois ordered phase 4 re-openings on June 26. Another critical element missing in our model corresponds to preferential shelter-in-place per age group. Even when mortality appears to more strongly impact the elderly, local mortality is low. This may be due to higher compliance of that population with shelter-at-home and other sanitary measures, including wearing masks in public. Variable viral loads per disease stage [54] are missing in our model, but these are harder to calibrate due to the biosafety and time elements involved in quantitative PCR-RT. Nevertheless, we do foresee situations where this may be possible and pertinent. Finally, our model assumes agents have an effectively infinite memory to remember whom they have had contact with. Contact tracing has a stringent limit when performed manually, which can be expanded greatly by means of various information technologies. Hence, our model is not realistic in this sense, since it does not distinguish between these two cases: when an epidemic has reached certain critical mass, active cases will be underestimated.

Computationally, the Epidemiology Workbench is limited by the lack of true concurrency in Mesa, thereby impacting scaling properties of our simulation. Distributing the agents across multiple compute nodes in Python requires architectural changes in Mesa beyond the scope of our work. At present, efforts are under way to develop an Elixir-based ABM platform capable of addressing this limitation as part of the SPEC collaboration in the Computational Social Science community. Another critical bottleneck is random number generation, for which various strategies may be applied, including the use of (approximately) irrational numbers as coefficients of Fourier series.

A final aspect underscored by our research, in particular in the context of COVID-19, is the need for anticipatory mechanisms driving public policy measures. In this sense, simulation methods such as the one presented here or others lack convenience when introducing external events and the execution cost of recomputing complex models, and appear to make sense only at early stages of a epidemic process: once the disease spread has reached its exponential phase, the need moves from prediction to probabilistically qualified estimations of short term measure effectiveness. This suggests a different class of stochastic methods that not only predict expected trends but also recommend measures based on their effectiveness, similar to those used in high-speed trading of financial derivatives and futures. The complexity of the socio-technical character of the global economy and society demands these more powerful methods to successfully address the wicked character of a pandemic, in particular this one.

As of now, our efforts concentrate on seeking viable ways to package and deploy the Epidemiology Workbench across various cyberinfrastructure resources in order to make it available to other small cities (including training resources) and, particularly, to communities with strong presence of underrepresented minorities whose public health planning resources are heavily constrained.

## Data Availability

The authors have provided all code and computer experiment data online at a public GitHub repository, including the script to setup a cloud computing instance.

https://github.com/snunezcr/COVID19-mesa

## Acknowledgments

The authors wish to thank the following individuals for their contributions and support: Julie Pryde and Awais Vaid (Champaign-Urbana Public Health District), public health data and situational knowledge; Tomas de Camino Beck (ULead), epidemiology and mathematical biology; Carlos Rau’l Guti’errez-Guti’errez (ICANN/ISOC), economics and econometrics; Jacqueline Kazil and Tom Pike (Mesa/George Mason University), Mesa programming and Python multiprocessing; Rajesh Venkatachalapathy (Portland State University), Srikanth Mudigonda (Saint Louis University), Milton Friesen (Cardus, U Waterloo) and Jeff Graham (Susquehanna University) on agent-based modeling strategies (SPEC collaborative); Galen Arnold (NCSA UIUC), Python optimization strategies, and William Gropp and Jim Glasgow for institutional support (NCSA/UIUC). This work was supported by Illinois Informatics and the ACM/Intel SIGHPC Computational and Data Science Fellowship, 2017 cohort.

## Footnotes

1 See: https://github.com/snunezcr/COVID19-mesa

2 US Census Bureau population estimates for 2019. See: https://bit.ly/3hj1PnC

3 Idem. See: https://bit.ly/2WGcOjq

4 University of Illinois News Bureau. See: https://bit.ly/39jYj9K

5 See: https://bit.ly/3eOlISa

6 CDC COVID-19 Death Data and Resources. See: https://bit.ly/3hpap4A

7 State of Illinois. See: https://bit.ly/2WHU6I5

8 Idem. See: https://bit.ly/3hsxm6D

9 See: https://bit.ly/2BhCuv3

10 See: https://bit.ly/30wgS6M

11 See: https://bit.ly/3jvR2Zw

12 See: https://bit.ly/2BkMh3v

## References

[1] Mohammed S Abdo, Kamal Shah, Hanan A Wahash, and Satish K Panchal. On a comprehensive model of the novel coronavirus (covid-19) under mittag-leffler derivative. Chaos, Solitons &Fractals, page 109867, 2020.

[2] Ahmed S Abdulamir and Rand R Hafidh. The possible immunological pathways for the variable immunopathogenesis of covid–19 infections among healthy adults, elderly and children. Electronic Journal of General Medicine, 17(4), 2020.

[3] David Adam. Special report: The simulations driving the world’s response to COVID-19. Nature, 580(7803):316, 2020.

[4] Jacob B Aguilar, Jeremy Samuel Faust, Lauren M Westafer, and Juan B Gutierrez. Investigating the impact of asymptomatic carriers on covid-19 transmission. medRxiv, 2020.

[5] Marco Ajelli, Bruno Gonçalves, Duygu Balcan, Vittoria Colizza, Hao Hu, Jos’e J Ramasco, Stefano Merler, and Alessandro Vespignani. Comparing large-scale computational approaches to epidemic modeling: agent-based versus structured metapopulation models. BMC infectious diseases, 10(1):190, 2010.

[6] Jaffar A Al-Tawfiq. Asymptomatic coronavirus infection: Mers-cov and sars-cov-2 (covid-19). Travel Med Infect Dis, 27(101608):1–2, 2020.

[7] Titan M Alon, Matthias Doepke, Jane Olmstead-Rumsey, and Michele Tertilt. The impact of covid-19 on gender equality. Technical report, National Bureau of Economic Research, 2020.

[8] Governor G Ameh, Anuli Njoku, Jeff Inungu, and Mustafa Younis. Rural america and coronavirus epidemic: Challenges and solutions. European Journal of Environment and Public Health, 4(2):em0040, 2020.

[9] SM Arifin, Rumana Reaz Arifin, Dilkushi De Alwis Pitts, M Sohel Rahman, Sara Nowreen, Gregory R Madey, and Frank H Collins. Landscape epidemiology modeling using an agent-based model and a geographic information system. Land, 4(2):378–412, 2015.

[10] Benjamin Armbruster and Ekkehard Beck. Elementary proof of convergence to the mean-field model for the sir process. Journal of mathematical biology, 75(2):327–339, 2017.

[11] Yan Bai, Lingsheng Yao, Tao Wei, Fei Tian, Dong-Yan Jin, Lijuan Chen, and Meiyun Wang. Presumed asymptomatic carrier transmission of covid-19. Jama, 323(14):1406–1407, 2020.

[12] Scott R Baker, Nicholas Bloom, Steven J Davis, and Stephen J Terry. Covid-induced economic uncertainty. Technical report, National Bureau of Economic Research, 2020.

[13] V Balachandar, I Mahalaxmi, J Kaavya, G Vivekanandhan, S Ajithkumar, N Arul, G Singaravelu, N Senthil Kumar, and S Mohana Devi Covid-19: emerging protective measures. European Review for Medical and Pharmacological Sciences, 24(6):3422–3425, 2020.

[14] Linlin Bao, Wei Deng, Hong Gao, Chong Xiao, Jiayi Liu, Jing Xue, Qi Lv, Jiangning Liu, Pin Yu, Yanfeng Xu, et al. Lack of reinfection in rhesus macaques infected with sars-cov-2. bioRxiv, 2020.

[15] Jean-Noel Barrot, Basile Grassi, and Julien Sauvagnat. Sectoral effects of social distancing. Available at SSRN, 2020.

[16] Maurice S Bartlett. Deterministic and stochastic models for recurrent epidemics. In Proceedings of the third Berkeley symposium on mathematical statistics and probability, volume 4(81), page 109, 1956.

[17] Martin Bicher and Niki Popper. Agent-based derivation of the sir-differential equations. In 2013 8th EUROSIM Congress on Modelling and Simulation, pages 306–311. IEEE, 2013.

[18] James RM Black, Chris Bailey, and Charles Swanton. Covid-19: the case for health-care worker screening to prevent hospital transmission. The Lancet, 2020.

[19] Konstantin B Blyuss and Yuliya N Kyrychko. Effects of latency and age structure on the dynamics and containment of covid-19. medRxiv, 2020.

[20] Eric Bonabeau. Agent-based modeling: Methods and techniques for simulating human systems. Proceedings of the national academy of sciences, 99(suppl 3):7280–7287, 2002.

[21] Alberto Botta, Eugenio Caverzasi, and Alberto Russo. Fighting the covid-19 emergency and relaunching the european economy: debt monetization and recovery bonds, 2020.

[22] Spiros Bougheas. The economy on ice: Meeting the economic challenges during and after the covid-19 crisis. Available at SSRN 3563536, 2020.

[23] Maged N Kamel Boulos and Estella M Geraghty. Geographical tracking and mapping of coronavirus disease covid-19/severe acute respiratory syndrome coronavirus 2 (sars-cov-2) epidemic and associated events around the world: how 21st century gis technologies are supporting the global fight against outbreaks and epidemics, 2020.

[24] Lydia Bourouiba. Turbulent gas clouds and respiratory pathogen emissions: potential implications for reducing transmission of covid-19. Jama, 2020.

[25] Francisco Buera, Roberto Fattal-Jaef, A Neumeyer, and Yongseok Shin. The economic ripple effects of covid-19. Unpublished manuscript. Available at the World Bank Development Policy and COVID-19— eSeminar Series, 2020.

[26] Yanan Cao, Lin Li, Zhimin Feng, Shengqing Wan, Peide Huang, Xiaohui Sun, Fang Wen, Xuanlin Huang, Guang Ning, and Weiqing Wang. Comparative genetic analysis of the novel coronavirus (2019-ncov/sars-cov-2) receptor ace2 in different populations. Cell Discovery, 6(1):1–4, 2020.

[27] Anindya S Chakrabarti and Bikas K Chakrabarti. Statistical theories of income and wealth distribution. Economics: The Open-Access, Open-Assessment E-Journal, 4(2010-4):1–31, 2010.

[28] Fabio ACC Chalub and Max O Souza. The sir epidemic model from a pde point of view. Mathematical and Computer Modelling, 53(7-8):1568–1574, 2011.

[29] Sung Nok Chiu, Dietrich Stoyan, Wilfrid S Kendall, and Joseph Mecke. Stochastic geometry and its applications. John Wiley &Sons, 2013.

[30] Olivier Coibion, Yuriy Gorodnichenko, and Michael Weber. Labor markets during the covid-19 crisis: A preliminary view. Technical report, National Bureau of Economic Research, 2020.

[31] Lucian Gideon Conway Iii, Shailee R Woodard, and Alivia Zubrod. Social psychological measurements of covid-19: Coronavirus perceived threat, government response, impacts, and experiences questionnaires, 2020.

[32] Nicholas G Davies, Petra Klepac, Yang Liu, Kiesha Prem, Mark Jit, Rosalind M Eggo, CMMID COVID-19 working group, et al. Age-dependent effects in the transmission and control of covid-19 epidemics. medRxiv, 2020.

[33] Michael Day. Covid-19: identifying and isolating asymptomatic people helped eliminate virus in Italian village. Bmj, 368:m1165, 2020.

[34] Carlos Del Rio and Preeti N Malani. Covid-19—new insights on a rapidly changing epidemic. Jama, 323(14):1339–1340, 2020.

[35] Ramses Djidjou-Demasse, Yannis Michalakis, Marc Choisy, Micea T Sofonea, and Samuel Alizon. Optimal covid-19 epidemic control until vaccine deployment. medRxiv, 2020.

[36] Marco D’Orazio, Gabriele Bernardini, and Enrico Quagliarini. How to restart? an agent-based simulation model towards the definition of strategies for covid-19” second phase” in public buildings. arXiv preprint 2004.12927, 2020.

[37] Jennifer Beam Dowd, Liliana Andriano, David M Brazel, Valentina Rotondi, Per Block, Xuejie Ding, Yan Liu, and Melinda C Mills. Demographic science aids in understanding the spread and fatality rates of covid-19. Proceedings of the National Academy of Sciences, 117(18):9696–9698, 2020.

[38] David M Eddy, William Hollingworth, J Jaime Caro, Joel Tsevat, Kathryn M McDonald, and John B Wong. Model transparency and validation: a report of the ISPOR-SMDM Modeling Good Research Practices Task Force–7. Medical Decision Making, 32(5):733–743, 2012.

[39] Ezekiel J Emanuel, Govind Persad, Ross Upshur, Beatriz Thome, Michael Parker, Aaron Glickman, Cathy Zhang, Connor Boyle, Maxwell Smith, and James P Phillips. Fair allocation of scarce medical resources in the time of covid-19, 2020.

[40] Jim AC Everett, Clara Colombatto, Vladimir Chituc, William J Brady, and Molly Crockett. The effectiveness of moral messages on public health behavioral intentions during the covid-19 pandemic, 2020.

[41] Neil Ferguson, Daniel Laydon, Gemma Nedjati Gilani, Natsuko Imai, Kylie Ainslie, Marc Baguelin, Sangeeta Bhatia, Adhiratha Boonyasiri, ZULMA Cucunuba Perez, Gina Cuomo-Dannenburg, et al. Report 9: Impact of non-pharmaceutical interventions (npis) to reduce covid19 mortality and healthcare demand, 2020.

[42] Dale Fisher and Annelies Wilder-Smith. The global community needs to swiftly ramp up the response to contain covid-19. The Lancet, 395(10230):1109–1110, 2020.

[43] Monica Gandhi, Deborah S Yokoe, and Diane V Havlir. Asymptomatic transmission, the achilles’ heel of current strategies to control covid-19, 2020.

[44] Ugo Gentilini, Mohamed Almenfi, Ian Orton, and P Dale. Social protection and jobs responses to covid-19: a real-time review of country measures. Live Document. World Bank, Washington, DC. http://www.ugogentilini.net/wp-content/uploads/2020/03/global-review-of-social-protection-responsesto-COVID-19-2.pdf, 2020.

[45] Rafael Gonz’alez-Val. Us city-size distribution and space. Spatial Economic Analysis, 14(3):283–300, 2019.

[46] Kenneth W Goodman and Eric M Meslin. Ethics, information technology, and public health: duties and challenges in computational epidemiology. In Public Health Informatics and Information Systems, pages 191–209. Springer, 2014.

[47] Trisha Greenhalgh, Manuel B Schmid, Thomas Czypionka, Dirk Bassler, and Laurence Gruer. Face masks for the public during the covid-19 crisis. Bmj, 369, 2020.

[48] Ellina Grigorieva, Evgenii Khailov, and Andrei Korobeinikov. Optimal quarantine strategies for covid-19 control models. arXiv preprint 2004.10614, 2020.

[49] Daniel F Gudbjartsson, Agnar Helgason, Hakon Jonsson, Olafur T Magnusson, Pall Melsted, Gud-mundur L Norddahl, Jona Saemundsdottir, Asgeir Sigurdsson, Patrick Sulem, Arna B Agustsdottir, et al. Spread of sars-cov-2 in the icelandic population. New England Journal of Medicine, 2020.

[50] Thomas Hale, Anna Petherick, Toby Phillips, and Samuel Webster. Variation in government responses to covid-19. Blavatnik School of Government Working Paper, 31, 2020.

[51] M Elizabeth Halloran, Neil M Ferguson, Stephen Eubank, Ira M Longini, Derek AT Cummings, Bryan Lewis, Shufu Xu, Christophe Fraser, Anil Vullikanti, Timothy C Germann, et al. Modeling targeted layered containment of an influenza pandemic in the united states. Proceedings of the National Academy of Sciences, 105(12):4639–4644, 2008.

[52] Bruce Hannon. Ecological pricing and economic efficiency. Ecological Economics, 36(1):19–30, 2001.

[53] Johannes Haushofer and JCE Metcalf. Combining behavioral economics and infectious disease epidemiology to mitigate the covid-19 outbreak. Princeton University, March, 6, 2020.

[54] Xi He, Eric HY Lau, Peng Wu, Xilong Deng, Jian Wang, Xinxin Hao, Yiu Chung Lau, Jessica Y Wong, Yujuan Guan, Xinghua Tan, et al. Temporal dynamics in viral shedding and transmissibility of covid-19. Nature medicine, 26(5):672–675, 2020.

[55] Brian W Head et al. Wicked problems in public policy. Public policy, 3(2):101, 2008.

[56] Christopher I Jarvis, Kevin Van Zandvoort, Amy Gimma, Kiesha Prem, Petra Klepac, G James Rubin, and W John Edmunds. Quantifying the impact of physical distance measures on the transmission of covid-19 in the uk. BMC medicine, 18(1):1–10, 2020.

[57] Chunyan Ji and Daqing Jiang. Threshold behaviour of a stochastic sir model. Applied Mathematical Modelling, 38(21-22):5067–5079, 2014.

[58] Kenneth M Johnson. An older population increases estimated covid-19 death rates in rural america, 2020.

[59] Frank Kachanoff, Yochanan Bigman, Kyra Kapsaskis, and Kurt Gray. Measuring two distinct psychological threats of covid-19 and their unique impacts on wellbeing and adherence to public health behaviors, 2020.

[60] Nirmal Kandel, Stella Chungong, Abbas Omaar, and Jun Xing. Health security capacities in the context of covid-19 outbreak: an analysis of international health regulations annual report data from 182 countries. The Lancet, 2020.

[61] Stephen M Kissler, Christine Tedijanto, Edward Goldstein, Yonatan H Grad, and Marc Lipsitch. Projecting the transmission dynamics of sars-cov-2 through the postpandemic period. Science, 2020.

[62] Abhiteja Konda, Abhinav Prakash, Gregory A Moss, Michael Schmoldt, Gregory D Grant, and Supratik Guha. Aerosol filtration efficiency of common fabrics used in respiratory cloth masks. ACS nano, 2020.

[63] Jeffrey E Kottemann, Fred D Davis, and William E Remus. Computer-assisted decision making: Performance, beliefs, and the illusion of control. Organizational Behavior and Human Decision Processes, 57(1):26–37, 1994.

[64] Tobias Kretz and Michael Schreckenberg. Moore and more and symmetry. In Pedestrian and evacuation dynamics 2005, pages 297–308. Springer, 2007.

[65] Andr’e Kurmann, Etienne Lal’e, and Lien Ta. The impact of covid-19 on us employment and hours: Real-time estimates with homebase data. May). http://www.andrekurmann.com/hbcovid, 2020.

[66] Jan H Kwakkel, Warren E Walker, and Marjolijn Haasnoot. Coping with the wickedness of public policy problems: approaches for decision making under deep uncertainty, 2016.

[67] Shengjie Lai, Nick W Ruktanonchai, Liangcai Zhou, Olivia Prosper, Wei Luo, Jessica R Floyd, Amy Wesolowski, Mauricio Santillana, Chi Zhang, Xiangjun Du, et al. Effect of non-pharmaceutical interventions to contain covid-19 in china, 2020.

[68] Siu Kwan Lam, Antoine Pitrou, and Stanley Seibert. Numba: A llvm-based python jit compiler. In Proceedings of the Second Workshop on the LLVM Compiler Infrastructure in HPC, pages 1–6, 2015.

[69] Stephen A Lauer, Kyra H Grantz, Qifang Bi, Forrest K Jones, Qulu Zheng, Hannah R Meredith, Andrew S Azman, Nicholas G Reich, and Justin Lessler. The incubation period of coronavirus disease 2019 (covid-19) from publicly reported confirmed cases: estimation and application. Annals of internal medicine, 2020.

[70] Ju Sung Lee, Tatiana Filatova, Arika Ligmann-Zielinska, Behrooz Hassani-Mahmooei, Forrest Stonedahl, Iris Lorscheid, Alexey Voinov, Gary Polhill, Zhanli Sun, and Dawn Cassandra Parker. The complexities of agent-based modeling output analysis. The journal of artificial societies and social simulation, 18(4), 2015.

[71] Joseph A Lewnard and Nathan C Lo. Scientific and ethical basis for social-distancing interventions against covid-19. The Lancet. Infectious diseases, 2020.

[72] Yuguo Lin, Daqing Jiang, and Peiyan Xia. Long-time behavior of a stochastic sir model. Applied Mathematics and Computation, 236:1–9, 2014.

[73] JA Machado, Maria Eug’enia Mata, and Ant’onio M Lopes. Fractional state space analysis of economic systems. Entropy, 17(8):5402–5421, 2015.

[74] Christos Makridis and Jonathan Hartley. The cost of covid-19: A rough estimate of the 2020 us gdp impact. Special Edition Policy Brief, 2020.

[75] Oded Maler, A’d’am M Hal’asz, Olivier Lebeltel, and Ouri Maler. Exploring synthetic mass action models. In International Workshop on Hybrid Systems Biology, pages 97–110. Springer, 2014.

[76] Lenore Manderson and Susan Levine. Covid-19, risk, fear, and fall-out, 2020.

[77] David Masad and Jacqueline Kazil. Mesa: an agent-based modeling framework. In 14th PYTHON in Science Conference, pages 53–60, 2015.

[78] Warwick McKibbin and Roshen Fernando. 3 the economic impact of covid-19. Economics in the Time of COVID-19, page 45, 2020.

[79] Warwick J McKibbin and Roshen Fernando. The global macroeconomic impacts of covid-19: Seven scenarios. 2020.

[80] Joseph S Meisel. Air raid shelter policy and its critics in britain before the second world war. Twentieth Century British History, 5(3):300–319, 1994.

[81] Timo Meynhardt. Public value inside: What is public value creation? Intl Journal of Public Administration, 32(3-4):192–219, 2009.

[82] Aaron Miller, Mac Josh Reandelar, Kimberly Fasciglione, Violeta Roumenova, Yan Li, and Gonzalo H Otazu. Correlation between universal bcg vaccination policy and reduced morbidity and mortality for covid-19: an epidemiological study. MedRxiv, 2020.

[83] Mukesh Kumar Mishra. The world after covid-19 and its impact on global economy. 2020.

[84] Wesley C Mitchell. The quantity theory of the value of money. Journal of Political Economy, 4(2):139– 165, 1896.

[85] Kenji Mizumoto, Katsushi Kagaya, Alexander Zarebski, and Gerardo Chowell. Estimating the asymptomatic proportion of coronavirus disease 2019 (covid-19) cases on board the diamond princess cruise ship, yokohama, japan, 2020. Eurosurveillance, 25(10):2000180, 2020.

[86] Seyed M Moghadas, Affan Shoukat, Meagan C Fitzpatrick, Chad R Wells, Pratha Sah, Abhishek Pandey, Jeffrey D Sachs, Zheng Wang, Lauren A Meyers, Burton H Singer, et al. Projecting hospital utilization during the covid-19 outbreaks in the united states. Proceedings of the National Academy of Sciences, 117(16):9122–9126, 2020.

[87] Stephen E Moore and Eric Okyere. Controlling the transmission dynamics of covid-19. arXiv preprint 2004.00443, 2020.

[88] L Nirenberg. Generalized degree and nonlinear problems. In Contributions to nonlinear functional analysis, pages 1–9. Elsevier, 1971.

[89] Barbara Nussbaumer-Streit, Verena Mayr, Andreea Iulia Dobrescu, Andrea Chapman, Emma Persad, Irma Klerings, Gernot Wagner, Uwe Siebert, Claudia Christof, Casey Zachariah, et al. Quarantine alone or in combination with other public health measures to control covid-19: a rapid review. Cochrane Database of Systematic Reviews, (4), 2020.

[90] Saad B Omer, Preeti Malani, and Carlos Del Rio. The covid-19 pandemic in the us: a clinical update. JAMA, 2020.

[91] World Health Organization et al. Laboratory testing for coronavirus disease (covid-19) in suspected human cases: interim guidance, 19 march 2020. Technical report, World Health Organization, 2020.

[92] Miyo Ota. Will we see protection or reinfection in covid-19?, 2020.

[93] Henrïette S Otter, Anne van der Veen, and Huib J de Vriend. Abloom: Location behaviour, spatial patterns, and agent-based modelling. Journal of Artificial Societies and Social Simulation, 4(4), 2001.

[94] Jason Phua, Li Weng, Lowell Ling, Moritoki Egi, Chae-Man Lim, Jigeeshu Vasishtha Divatia, Babu Raja Shrestha, Yaseen M Arabi, Jensen Ng, Charles D Gomersall, et al. Intensive care management of coronavirus disease 2019 (covid-19): challenges and recommendations. The Lancet Respiratory Medicine, 2020.

[95] Janice Probst, Jan Marie Eberth, and Elizabeth Crouch. Structural urbanism contributes to poorer health outcomes for rural america. Health Affairs, 38(12):1976–1984, 2019.

[96] Hazhir Rahmandad and John Sterman. Heterogeneity and network structure in the dynamics of diffusion: Comparing agent-based and differential equation models. Management Science, 54(5):998– 1014, 2008.

[97] Carlos Romero-Rivas and Sara Rodr’iguez-Cuadrado. Moral decision-making and mental health during the covid-19 pandemic. 2020.

[98] Timothy W Russell, Joel Hellewell, Christopher I Jarvis, Kevin Van Zandvoort, Sam Abbott, Ruwan Ratnayake, Stefan Flasche, Rosalind M Eggo, W John Edmunds, Adam J Kucharski, et al. Estimating the infection and case fatality ratio for coronavirus disease (covid-19) using age-adjusted data from the outbreak on the diamond princess cruise ship, february 2020. Eurosurveillance, 25(12):2000256, 2020.

[99] Marcel Salath’e, Christian L Althaus, Richard Neher, Silvia Stringhini, Emma Hodcroft, Jacques Fellay, Marcel Zwahlen, Gabriela Senti, Manuel Battegay, Annelies Wilder-Smith, et al. Covid-19 epidemic in switzerland: on the importance of testing, contact tracing and isolation. Swiss medical weekly, 150(11-12):w20225, 2020.

[100] Junkichi Satsuma, R Willox, A Ramani, B Grammaticos, and AS Carstea. Extending the sir epidemic model. Physica A: Statistical Mechanics and its Applications, 336(3-4):369–375, 2004.

[101] Günter Schneckenreither, Nikolas Popper, Günther Zauner, and Felix Breitenecker. Modelling sir-type epidemics by odes, pdes, difference equations and cellular automata–a comparative study. Simulation Modelling Practice and Theory, 16(8):1014–1023, 2008.

[102] Dag Sverre Seljebotn. Fast numerical computations with cython. In Proceedings of the 8th Python in Science Conference, volume 37, 2009.

[103] Yufang Shi, Ying Wang, Changshun Shao, Jianan Huang, Jianhe Gan, Xiaoping Huang, Enrico Bucci, Mauro Piacentini, Giuseppe Ippolito, and Gerry Melino. Covid-19 infection: the perspectives on immune responses, 2020.

[104] Gitanjali R Shinde, Asmita B Kalamkar, Parikshit N Mahalle, Nilanjan Dey, Jyotismita Chaki, and Aboul Ella Hassanien. Forecasting models for coronavirus disease (covid-19): A survey of the state-of-the-art. SN Computer Science, 1(4):1–15, 2020.

[105] Constantinos I Siettos and Lucia Russo. Mathematical modeling of infectious disease dynamics. Virulence, 4(4):295–306, 2013.

[106] Henrik Sjödin, Annelies Wilder-Smith, Sarah Osman, Zia Farooq, and Joacim Rocklöv. Only strict quarantine measures can curb the coronavirus disease (covid-19) outbreak in italy, 2020. Eurosurveillance, 25(13):2000280, 2020.

[107] Jiumeng Sun, Wan-Ting He, Lifang Wang, Alexander Lai, Xiang Ji, Xiaofeng Zhai, Gairu Li, Marc A Suchard, Jin Tian, Jiyong Zhou, et al. Covid-19: epidemiology, evolution, and cross-disciplinary perspectives. Trends in Molecular Medicine, 2020.

[108] Vasily E Tarasov and Valentina V Tarasova. Long and short memory in economics: fractional-order difference and differentiation. arXiv preprint 1612.07903, 2016.

[109] In’es Tejado, Duarte Val’erio, and Nuno Val’erio. Fractional calculus in economic growth modeling. the portuguese case. In ICFDA’14 International Conference on Fractional Differentiation and Its Applications 2014, pages 1–6. IEEE, 2014.

[110] Linda Thunström, Stephen C Newbold, David Finnoff, Madison Ashworth, and Jason F Shogren. The benefits and costs of using social distancing to flatten the curve for covid-19. Journal of Benefit-Cost Analysis, pages 1–27, 2020.

[111] Bruno Tilocca, Alessio Soggiu, Vincenzo Musella, Domenico Britti, Maurizio Sanguinetti, Andrea Urbani, and Paola Roncada. Molecular basis of covid-19 relationships in different species: a one health perspective. Microbes and Infection, 2020.

[112] Alexis Akira Toda. Susceptible-infected-recovered (sir) dynamics of covid-19 and economic impact. arXiv preprint 2003.11221, 2020.

[113] Elisabetta Tornatore, Stefania Maria Buccellato, and Pasquale Vetro. Stability of a stochastic sir system. Physica A: Statistical Mechanics and its Applications, 354:111–126, 2005.

[114] Pauli Virtanen, Ralf Gommers, Travis E Oliphant, Matt Haberland, Tyler Reddy, David Cournapeau, Evgeni Burovski, Pearu Peterson, Warren Weckesser, Jonathan Bright, et al. Scipy 1.0: fundamental algorithms for scientific computing in python. Nature methods, 17(3):261–272, 2020.

[115] M Wang, Z Hu, J Liu, P Pang, G Fu, A Qian, S Chen, L Lin, G Cao, H Sun, et al. Positive rt-pcr test results in discharged covid-19 patients: Reinfection or residual?, 2020.

[116] Annelies Wilder-Smith, Calvin J Chiew, and Vernon J Lee. Can we contain the covid-19 outbreak with the same measures as for sars? The Lancet Infectious Diseases, 2020.

[117] Chufen Wu, Yong Yang, Qianyi Zhao, Yanling Tian, and Zhiting Xu. Epidemic waves of a spatial sir model in combination with random dispersal and non-local dispersal. Applied Mathematics and Computation, 313:122–143, 2017.

[118] Joseph T Wu, Kathy Leung, Mary Bushman, Nishant Kishore, Rene Niehus, Pablo M de Salazar, Benjamin J Cowling, Marc Lipsitch, and Gabriel M Leung. Estimating clinical severity of covid-19 from the transmission dynamics in wuhan, china. Nature Medicine, pages 1–5, 2020.

[119] Wenlei Xiao, Qiang Liu, J Huan, Pengpeng Sun, Liuquan Wang, Chenxin Zang, Sanying Zhu, and Liansheng Gao. A cybernetics-based dynamic infection model for analyzing sars-cov-2 infection stability and predicting uncontrollable risks. medRxiv, 2020.

[120] Yonghong Xiao and Mili Estee Torok. Taking the right measures to control covid-19. The Lancet Infectious Diseases, 2020.

[121] Yuri Yegorov. Role of density and field in spatial economics. Contemporary Issues in Urban and Regional Economics. NY: Nova Science Publishers, pages 55–78, 2005.

[122] Xingxia Yu and Rongrong Yang. Covid-19 transmission through asymptomatic carriers is a challenge to containment. Influenza and Other Respiratory Viruses, 2020.

[123] Xianghua Zhang and Ke Wang. Stochastic sir model with jumps. Applied Mathematics Letters, 26(8):867–874, 2013.

